# Bi-allelic loss-of-function variants in *PPFIBP1* cause a neurodevelopmental disorder with microcephaly, epilepsy and periventricular calcifications

**DOI:** 10.1101/2022.04.04.22273309

**Authors:** Erik Rosenhahn, Thomas J. O’Brien, Maha S. Zaki, Ina Sorge, Dagmar Wieczorek, Kevin Rostasy, Antonio Vitobello, Sophie Nambot, Fowzan S. Alkuraya, Mais O. Hashem, Amal Alhashem, Brahim Tabarki, Abdullah S. Alamri, Ayat H. Al Safar, Dalal K. Bubshait, Nada F. Alahmady, Joseph G. Gleeson, Mohamed S. Abdel-Hamid, Nicole Lesko, Sofia Ygberg, Sandrina P. Correia, Anna Wredenberg, Shahryar Alavi, Seyed M. Seyedhassani, Mahya Ebrahimi Nasab, Haytham Hussien, Tarek Omar, Ines Harzallah, Renaud Touraine, Homa Tajsharghi, Heba Morsy, Henry Houlden, Mohammad Shahrooei, Maryam Ghavideldarestani, Johannes R. Lemke, Heinrich Sticht, Rami Abou Jamra, Andre E. X. Brown, Reza Maroofian, Konrad Platzer

## Abstract

*PPFIBP1* encodes for the liprin-β1 protein which has been shown to play a role in neuronal outgrowth and synapse formation in *Drosophila melanogaster*. By exome sequencing, we detected nine ultra-rare homozygous loss-of-function variants in 14 individuals from 10 unrelated families. The individuals presented with moderate to profound developmental delay, often refractory early-onset epilepsy and progressive microcephaly. Further common clinical findings included muscular hypertonia, spasticity, failure to thrive and short stature, feeding difficulties, impaired hearing and vision, and congenital heart defects. Neuroimaging revealed abnormalities of brain morphology with leukoencephalopathy, cortical abnormalities, and intracranial periventricular calcifications as major features. In a fetus with intracranial calcifications, we identified a rare homozygous missense variant that by structural analysis was predicted to disturb the topology of the SAM-domain region that is essential for protein-protein interaction. For further insight in the effects of *PPFIBP1* loss-of-function, we performed automated behavioural phenotyping of a *Caenorhabditis elegans PPFIBP1/hlb-1* knockout model which revealed defects in spontaneous and light-induced behaviour and confirmed resistance to the acetylcholinesterase inhibitor aldicarb suggesting a defect in the neuronal presynaptic zone. In conclusion, we present bi-allelic loss-of-function variants in *PPFIBP1* as a novel cause of an autosomal recessive neurodevelopmental disorder.

## Introduction

*PPFIBP1* (GenBank: NM_003622.4; MIM: 603141) encodes for the PPFIA binding protein 1, also known as liprin-β1. Liprin-β1 belongs to the liprin-protein family whose members are characterised by a highly conserved N-terminal coiled coil region and three adjacent C-terminal sterile alpha motifs (SAM domains) that form multiple protein binding surfaces and allow for protein-protein interaction.^1–3^ In mammals, the liprin family comprises four liprin-α proteins (liprin-α1-4) and two liprin-β proteins (liprin-β1 and -β2). Liprin-β1 has the ability to homodimerize and to heterodimerize with the homologous α-liprins.^1^ In addition, liprin-β1 and liprin-α1 co-localize to the cell membrane and to the periphery of focal adhesions in fibroblast cell cultures (COS cells).^1,4^ Liprin-α proteins are major scaffold proteins involved in synapse formation, synaptic signaling and axonal transport processes via the assembly of large protein complexes.^5,6^ Although yet to be confirmed, it has been suggested, that liprin-β1 could play a role in the regulation of liprin-α mediated protein assemblies.^1,6,7^ In line with this is the observation that liprin-β1 forms a ternary complex with liprin-α2 and CASK,^3^ a presynaptic scaffolding protein essential for neurodevelopment.^8,9^ A previous knock-out model of the *PPFIBP1* homolog *hlb-1* in *C. elegans* showed abnormal locomotion behavior. Furthermore, abnormal and decreased distribution of snb-1, an ortholog of human VAMP-family proteins involved in presynaptic vesicle release, increased presynaptic zone size and resistance to the acetylcholinesterase inhibitor aldicarb indicated a role of *hlb-1* in the regulation of presynaptic function.^10^ Pointing towards a role in neurodevelopment, null-allele mutants of the liprin-β1 orthologue liprin-β resulted in altered axon outgrowth and synapse formation of R7 photoreceptors and also reduced larval neuromuscular junction (NMJ) size in *D. melanogaster*.^7^ Indeed, *PPFIBP1* has been proposed as a candidate gene for congenital microcephaly based on a single family although this link remains tentative and requires independent confirmation.^10^

Here we describe a cohort of 14 individuals with a neurodevelopmental disorder from ten unrelated families harboring homozygous loss-of-function (LoF) variants and a fetus with a missense variant in *PPFIBP1*.

## Subjects and methods

### Patient recruitment and consent

The study was approved by the ethics committee of the University of Leipzig (402/16-ek). Written informed consent for molecular testing and permission for publication of the data was obtained from all individuals and/or their legal representatives by the referring physicians according to the guidelines of the ethics committees and institutional review boards of the respective institute.

All individuals were ascertained in the context of local diagnostic protocols followed by research evaluation of the sequencing data. The compilation of the cohort was supported by international collaboration and online matchmaking via GeneMatcher^11^ in the case of family 2, 4, 5, 6, 7, 8, 9, 10 and 11. Family 3 was recently published as part of a larger cohort of individuals with congenital microcephaly.^12^ In the context of our study we describe the phenotype in detail. Phenotypic and genotypic information were obtained from the referring collaborators using a standardized questionnaire.

### Exome sequencing

Trio exome sequencing (ES) and trio genome sequencing were performed for the affected individuals and the parents in the families 1, 2, 5 and 11 and in family 7, respectively. Singleton exome sequencing was performed for all affected individuals of the families 3, 4, 6, 8, 9 and 10 (see Supplemental methods for further details).

### Variant prioritization

We first analysed single nucleotide variants annotated in local and public mutation databases, as well as rare (MAF < 1%) potentially protein-damaging variants in known disease-associated genes (e.g. by using in silico panels like MorbidGenes^13^). Variants were prioritised based on the mode of inheritance, impact on the gene product, minor allele frequency and *in silico* predicted pathogenicity. Since no pathogenic or likely pathogenic variants in known disease genes could be found and the families gave consent for further research, we then evaluated the sequencing data in a research setting aiming to identify potentially causative variants in novel candidate genes. For this purpose, variants in potential candidate genes were prioritised according to the parameters described above and by considering further data such as mutational constraint parameters from gnomAD^14^ or expression patterns in GTEx.^15^

All variants in *PPFIBP1* described here were aligned to the human reference genome version GRCh38 (hg38) and to the transcript NM_003622.4 (Ensembl: ENST00000228425.11) representing the transcript with the highest expression across all tissues^15^ and the MANE Select v0.95 default transcript. The pathogenicity of all described variants was classified according to the guidelines of the American College of Medical Genetics (ACMG)^16^ (Table S1).

### Structural analysis

Structural analysis of the p.(Gly726Val) variant was performed based on the crystal structure of murine PPFIBP1 (PDB: 3TAD ^3^), which exhibits 97% sequence identity to its human ortholog in the region of the SAM domains. The exchange was modeled with SwissModel^17^ and RasMol^18^ was used for structure analysis and visualization. Structural analysis for the other variants was not necessary as all are LoF variants and predicted to lead to a loss of protein.

### Mutant C. elegans generation

The knockout worm model was designed and made by SunyBiotech (Fuzhou, Fujian, China) in their reference N2 background. CRISPR guide RNA was designed to target a large deletion (17118 bp) starting close to the start codon and excising all exons from the gene. Deletions were confirmed by PCR.

### Worm preparation

All strains were cultured on Nematode Growth Medium at 20°C and fed with *E. coli* (OP50) following standard procedure.^19^ Synchronised populations of young adult worms for imaging were cultured by bleaching unsynchronised gravid adults, and allowing L1 diapause progeny to develop for two and a half days at 20°C.^20^ On the day of imaging, young adults were washed in M9^21^, transferred to the imaging plates (3 worms per well) using a COPAS 500 Flow Pilot^22^, and returned to a 20°C incubator for 3.5 hours. Plates were then transferred onto the multi-camera tracker for another 30 minutes to habituate prior to imaging^23^. For drug experiments, imaging plates were dosed with the compound at the desired concentration one day prior to imaging. Worms were then dispensed and tracked as described above, except for the 1h exposure time where worms were returned to a 20°C incubator for 30 minutes and then transferred to the tracker for 30 minutes prior to imaging^24^.

### Image acquisition, processing and feature extraction

Videos were acquired and processed following methods previously described in detail^25^. Briefly, videos were acquired in a room with a nominal temperature of 20°C at 25 frames per second and a resolution of 12.4 µm px^-1^. Three videos were taken sequentially: a 5-minute pre-stimulus video, a 6-minute blue light recording with three 10-second blue light pulses starting at 60, 160 and 260 seconds, and a 5-minute post-stimulus recording.

Videos were segmented and tracked using Tierpsy Tracker.^24^ After segmentation and skeletonisation, a manual threshold was applied to filter skeletonised objects, likely to be non-worms from feature extraction, that did not meet the following criteria: 200 – 2000 µM length, 20 – 500 µM width. Tierpsy Tracker’s viewer was also used to mark wells with visible contamination, agar damage, or excess liquid as “bad”, and exclude these wells from downstream analysis.

Following tracking, we extracted a previously-defined set of 3076 behavioural features for each well in each of the three videos (pre-stimulus, blue light and post-stimulus).^26^ The extraction of behavioural features was performed on a per-track basis and are then averaged across tracks to produce a single feature vector for each well. Statistically significant differences in the pre-stimulus, post-stimulus and blue-light behavioural feature sets extracted from the loss-of-function mutant compared to the N2 reference strain were calculated using block permutation t-tests (code available on GitHub, see web resources). Python (version 3.8.5) was used to perform the analysis, using n = 10^6^ permutations that were randomly shuffled within, but not between, the independent days of image acquisition in order to control for day-to-day variation in the experiments. The p-values were then corrected for multiple comparisons using the Benjamini-Hochberg procedure^27^ to control the false discovery rate at 5%.

### Pharyngeal pumping assay

Pharyngeal pumps per minute (ppm) of *C. elegans* strains were determined by counting grinder movements over a 15 second period by eye using a stereomicroscope^28^, n = 120 worms per strain. Grinder movements of a single worm were counted three times and the results recorded as an average of these values. Statistical differences in ppm between N2 reference strain and *hlb-1(syb4896)* were calculated using block permutation t-tests, using n = 10^3^ permutations.

## Results

### Clinical description

All individuals share a core phenotype of global developmental delay / intellectual disability (GDD/ID) and epilepsy. 13 were affected by profound or severe GDD/ID (13/14). They had not acquired speech (13/14) and showed impaired motor development (13/14). Most of them never achieved gross motor milestones such as sitting and walking except for individual 6-1 who was able to sit independently at the age of (removed due to medRxivpolicy) and individual 7 who could stand and walk. Individual 1 presented with moderate ID, developed expressive language skills and had a normal motor development. All individuals were affected by epilepsy: most commonly with focal seizures (9/14) including cases of impaired awareness (1/14) and focal to bilateral tonic-clonic (1/14) seizures. Furthermore, generalized onset seizures occurred in six of the individuals (6/14). Epileptic spasms were described in half of the cohort (7/14). Other reported seizure types included tonic (3/14) and myoclonic seizures (6/14). The median age of seizure onset was at two months with a range from the first day of life up to four years. Many individuals were initially affected by daily seizures (9/14). All individuals have been treated with multiple antiepileptic drugs (AED). However, seizures were refractory in six of the individuals (6/14). Seizures could be reasonably controlled in individual 1, individual 5-1, individual 6-1 and individual 8 with a therapy of Lamotrigine (LTG) and Oxcarbazepine (OXC), a therapy with Valproic acid (VPA), Clonazepam (CZP), Carbamazepine (CBZ), Levetiracetam (LEV) and LTG, a therapy with VPA, LEV and CZP and a therapy with Topiramate, VPA and LEV, respectively. The seizures completely ceased in individual 2, individual 7 and individual 10 under therapy with LEV and CZP, with VPA and with VPA and Diazepam, respectively. Individual 9 was reported to have had a seizure-free period. Electroencephalography (EEG) was performed in 12 individuals (Table S4). EEG findings included focal (3/12) or multifocal (5/12) interictal epileptiform discharges. In one individual, bilateral paroxysmal discharges were observed. Hypsarrhythmia was recorded in four individuals (4/12) who were also affected by epileptic spasms and thus met the criteria for West syndrome. In one of these cases, the phenotype progressed to Lennox-Gastaut syndrome later. All but individual 8 were affected by microcephaly, defined here by an occipitofrontal circumference (OFC) ≤ -2 standard deviations (SD) (range: <<-3 SD to -1.78 SD) at last assessment). The majority showed primary (9/14) and/or progressive (10/14) microcephaly. Secondary microcephaly developed in individual 1, individual 6-1 who had low OFC (−1.94 SD) already at birth, and individual 7. Individual 8 showed borderline low normal head circumference at the last assessment. Other common neurological findings comprise muscular hypertonia (10/14) up to spastic tetraplegia (8/14), but also muscular hypotonia (3/14), dystonic movements (2/14) and nystagmus (3/14).

Eight individuals were born small for gestational age (birthweight ≤ 10^th^ percentile; 8/14). Failure to thrive leading to decreased body weight (≤ -2SD) was seen in seven individuals (7/14) and short stature (height ≤ -2 SD) manifested in six (6/14). Some of the individuals exhibited feeding difficulties (6/14) and deglutition disorders were described in three of them.

Other repeatedly described symptoms include impaired hearing (4/14), ophthalmologic abnormalities (6/14), undescended testes (3/9) and congenital heart defects (6/14). The latter comprise patent ductus arteriosus (5/14, PDA), atrial septal defects (3/14, ASD), ventricular septal defects (2/14, VSD), a dilated left ventricle (1/14) and a coronary fistula (1/14) with mitral regurgitation and cardiomegaly. There were no overarching dysmorphic facial features in the affected individuals. (For an overview of the phenotypic spectrum see **Table 1** and **Figure 1A**. For further details on the phenotype of each individual see case reports in supplemental data and Table S4).

**Table 1.** Phenotypic and Genetic Features of Individuals with bi-allelic variants in PPFIBP1.

| Ind. | Age<sup>a</sup><br>(Sex) | Variant<br>(NM_003622.4) | Development | Seizure types<br>(age of onset) | MRI (age) | ICC<sup>b</sup> | Neurological<br>findings | Microce-<br>phaly | Growth | CHD | Hearing | Ocular<br>features |
| --- | --- | --- | --- | --- | --- | --- | --- | --- | --- | --- | --- | --- |
| Ind.<br>1 | (removed<br>due to<br>medRxiv<br>policy)<br>(M) | c.1146+1G>A,<br>p?, homozygous | moderate ID,<br>delayed<br>speech, normal<br>motor<br>development | focal impaired<br>awareness<br>(removed due to<br>medRxiv<br>policy) | normal (removed due to<br>medRxiv policy) | not<br>done | none | yes | SGA | no | normal | normal |
| Ind.<br>2 | (removed<br>due to<br>medRxiv<br>policy)<br>(F) | c.2654del,<br>p.(Tyr885Leufs*<br>4), homozygous | profound DD,<br>no speech,<br>unable to sit | focal,<br>generalized<br>tonic clonic<br>(removed due to<br>medRxiv<br>policy) | paucity of the WM,<br>ventriculomegaly,<br>hypoplastic CC, Blakes's<br>pouch cyst (removed due to<br>medRxiv policy) | yes | spastic<br>tetraplegia,<br>nystagmus | yes | short<br>stature,<br>low<br>weight | no | left<br>hypo-<br>acusis | bilateral<br>papillary<br>pallor, no eye<br>contact |
| Ind.<br>3-1 | (removed<br>due to<br>medRxiv<br>policy)<br>(M) | c.1368_1369del,<br>p.(Glu456Aspfs*<br>3), homozygous | profound DD,<br>no speech,<br>unable to sit | epileptic<br>spasms, focal,<br>tonic clonic,<br>tonic (removed<br>due to medRxiv<br>policy) | periventricular<br>leukomalacia, metopic<br>synostosis | yes | spastic<br>tetraplegia | yes | SGA,<br>low<br>weight | yes | impaired | poor fixation |
| Ind.<br>3-2 | (removed<br>due to<br>medRxiv<br>policy)<br>(M) | c.1368_1369del,<br>p.(Glu456Aspfs*<br>3), homozygous | profound DD,<br>no speech,<br>unable to sit | epileptic<br>spasms, LGS<br>(removed due to<br>medRxiv<br>policy) | moderate hyperintensity of<br>periventricular white matter,<br>mild ventriculomegaly<br>(removed due to medRxiv policy) | yes | spastic<br>tetraplegia | yes | SGA,<br>low<br>weight | yes | impaired | normal |
| Ind.<br>3-3 | (removed<br>due to<br>medRxiv<br>policy)<br>(M) | c.1368_1369del,<br>p.(Glu456Aspfs*<br>3), homozygous | profound DD,<br>no speech,<br>unable to sit | epileptic<br>spasms, focal,<br>multifocal<br>(removed due to<br>medRxiv<br>policy) | ventriculomegaly, abnormal<br>signal intensity of the WM,<br>bilateral temporal and left<br>occipital pachygyria (removed<br>due to medRxiv policy) | yes | spastic<br>tetraplegia | yes | SGA,<br>low<br>weight | yes | impaired | normal |
| Ind.<br>4 | (removed<br>due to<br>medRxiv<br>policy)<br>(M) | c.1368_1369del,<br>p.(Glu456Aspfs*<br>3), homozygous | profound DD,<br>no speech,<br>unable to sit | epileptic<br>spasms, focal,<br>generalized<br>tonic, status<br>epilepticus<br>(removed due to<br>medRxiv<br>policy) | ventriculomegaly, paucity of<br>the WM, bilateral parietal<br>and occipital pachygyria<br>(removed due to medRxiv policy) | yes | spastic<br>diplegia,<br>hyperreflexia | yes | SGA,<br>short<br>stature | yes | not<br>tested | haemorrhagic<br>retinitis,<br>chronic retinal<br>detachment,<br>right eye<br>exotropia w/<br>slow pupillary<br>reaction |
| Ind.<br>5-1 | (removed<br>due to<br>medRxiv<br>policy)<br>(F) | c.2413C>T,<br>p.(Arg805*),<br>homozygous | profound DD,<br>no speech,<br>unable to sit | focal,<br>myoclonic<br>(removed due to<br>medRxiv<br>policy) | not done | yes | spastic<br>tetraplegia | yes | SGA,<br>short<br>stature,<br>low<br>weight | no | normal | NA |

| Ind. | Age<sup>a</sup><br>(Sex) | Variant<br>(NM_003622.4) | Development | Seizure types<br>(age of onset) | MRI (age) | ICC<sup>b</sup> | Neurological<br>findings | Microce-<br>phaly | Growth | CHD | Hearing | Ocular<br>features |
| --- | --- | --- | --- | --- | --- | --- | --- | --- | --- | --- | --- | --- |
| Ind.<br>5-2 | removed<br>due to<br>medRxiv<br>policy (F) | c.2413C>T,<br>p.(Arg805*),<br>homozygous | profound DD,<br>no speech,<br>unable to sit | focal,<br>myoclonic,<br>tonic (removed<br>due to medRxiv<br>policy) | not done, ventriculomegaly<br>on CT | yes | hypertonia of<br>the limbs,<br>dystonia | yes | short<br>stature | no | normal | normal |
| Ind.<br>6-1 | removed<br>due to<br>medRxiv<br>policy (M) | c.1468C>T,<br>p.(Gln490*),<br>homozygous | profound DD,<br>no speech, sat<br>independently<br>at 6y | generalized<br>tonic clonic,<br>myoclonic<br>(removed due to<br>medRxiv<br>policy) | ventriculomegaly, cortical<br>atrophy, demyelination of<br>periventricular WM, thin<br>CC, cerebellar vermian<br>hypoplasia | not<br>done | hypertonia of<br>the limbs | yes | short<br>stature,<br>low<br>weight | no | normal | optic atrophy,<br>followed light |
| Ind.<br>6-2 | removed<br>due to<br>medRxiv<br>policy (M) | c.1468C>T,<br>p.(Gln490*),<br>homozygous | profound DD,<br>no speech, no<br>head support | generalized<br>tonic clonic,<br>myoclonic,<br>excessive<br>smacking<br>movements<br>(removed due to<br>medRxiv<br>policy) | asymmetrical<br>ventriculomegaly, cortical<br>atrophy, demyelination of<br>periventricular WM, thin<br>CC, cerebellar vermian<br>hypoplasia | yes | spasticity,<br>rigidity,<br>dystonic<br>movement | yes | short<br>stature,<br>low<br>weight | yes | normal | optic atrophy,<br>couldn't follow<br>light |
| Ind.<br>7 | removed<br>due to<br>medRxiv<br>policy (F) | c.403C>T,<br>p.(Arg135*),<br>homozygous | severe DD, no<br>speech, motor<br>delay but can<br>stand and walk | epileptic<br>spasms, focal<br>with apnoea,<br>myoclonic<br>(removed due to<br>medRxiv<br>policy) | normal at (removed due to<br>medRxiv policy); thin CC,<br>periventricular<br>dysmyelination, possibly<br>reduction of the WM at<br>(removed due to medRxiv<br>policy) | not<br>done | hypotonia | yes | normal | no | normal | normal |
| Ind.<br>8 | removed<br>due to<br>medRxiv<br>policy (F) | c.1417_1427del,<br>p.(Ala473Lysfs*<br>20),<br>homozygous | profound DD,<br>no speech,<br>unable to sit | generalized<br>tonic clonic<br>(removed due to<br>medRxiv<br>policy) | bilateral parietal pachygyria,<br>periventricular heterotopia,<br>ventriculomegaly,<br>hyperintensity and paucity<br>of the WM | NA | hypotonia,<br>nystagmus | no, but<br>low OFC | SGA | yes | normal | normal, but<br>poor fixation |
| Ind.<br>9 | removed<br>due to<br>medRxiv<br>policy (M) | c.1300C>T,<br>p.(Gln434*),<br>homozygous | severe DD, no<br>speech, can sit<br>but not walk | epileptic<br>spasms and<br>gaze (removed<br>due to medRxiv<br>policy) | abnormal | NA | spastic<br>tetraplegia, no<br>sphincter<br>control | yes | SGA | no | normal | blindness |

| Ind. | Age<sup>a</sup><br>(Sex) | Variant<br>(NM_003622.4) | Development | Seizure types<br>(age of onset) | MRI (age) | ICC<sup>b</sup> | Neurological<br>findings | Microce<br>phaly | Growth | CHD | Hearing | Ocular<br>features |
| --- | --- | --- | --- | --- | --- | --- | --- | --- | --- | --- | --- | --- |
| Ind.<br>10 | removed<br>due to<br>medRxiv<br>policy<br>(M) | c.2629C>T,<br>p.(Arg877*),<br>homozygous | severe DD, no<br>speech yet,<br>motor delay | focal<br>myoclonic,<br>epileptic<br>spasms<br>(removed due to<br>medRxiv<br>policy) | abnormal myelination of the<br>periventricular WM and at<br>corona radiata and centrum<br>semiovale, hypoplastic CC,<br>mild ventriculomegaly | NA | hypotonia,<br>nystagmus | yes | NA | no | normal | right ptosis,<br>left iris<br>coloboma,<br>diffuse<br>chorioretinal<br>degeneration |
| Fetus<br>(Ind.<br>11) | removed<br>due to<br>medRxiv<br>policy | c.2177G>T,<br>p.(Gly726Val),<br>homozygous | developmental<br>age estimated<br>around<br>(removed due to<br>medRxiv<br>policy) | - | - | Yes<sup>d</sup> | - | yes | IUGR | - | - | - |

Abbreviations: CC, corpus callosum; CHD, congenital heart defects; d, days; DD, developmental delay; F, female; IUGR, intrauterine growth restriction; GDD, global developmental delay; GW, gestational week; ICC, intracranial calcifications; ID, intellectual disability; LGS, Lennox-Gastaut syndrome; M, male; m, months; NA, not available; OFC, occipitofrontal circumference; SGA, small for gestational age; w, weeks; WM, white matter; y, years. Further clinical details are provided in Table S4.
<sup>a</sup> age at last assessment; <sup>b</sup> on CT scan; <sup>c</sup> deceased (age at death); <sup>d</sup> ICCs seen on x-ray babygram

**Figure 1.**
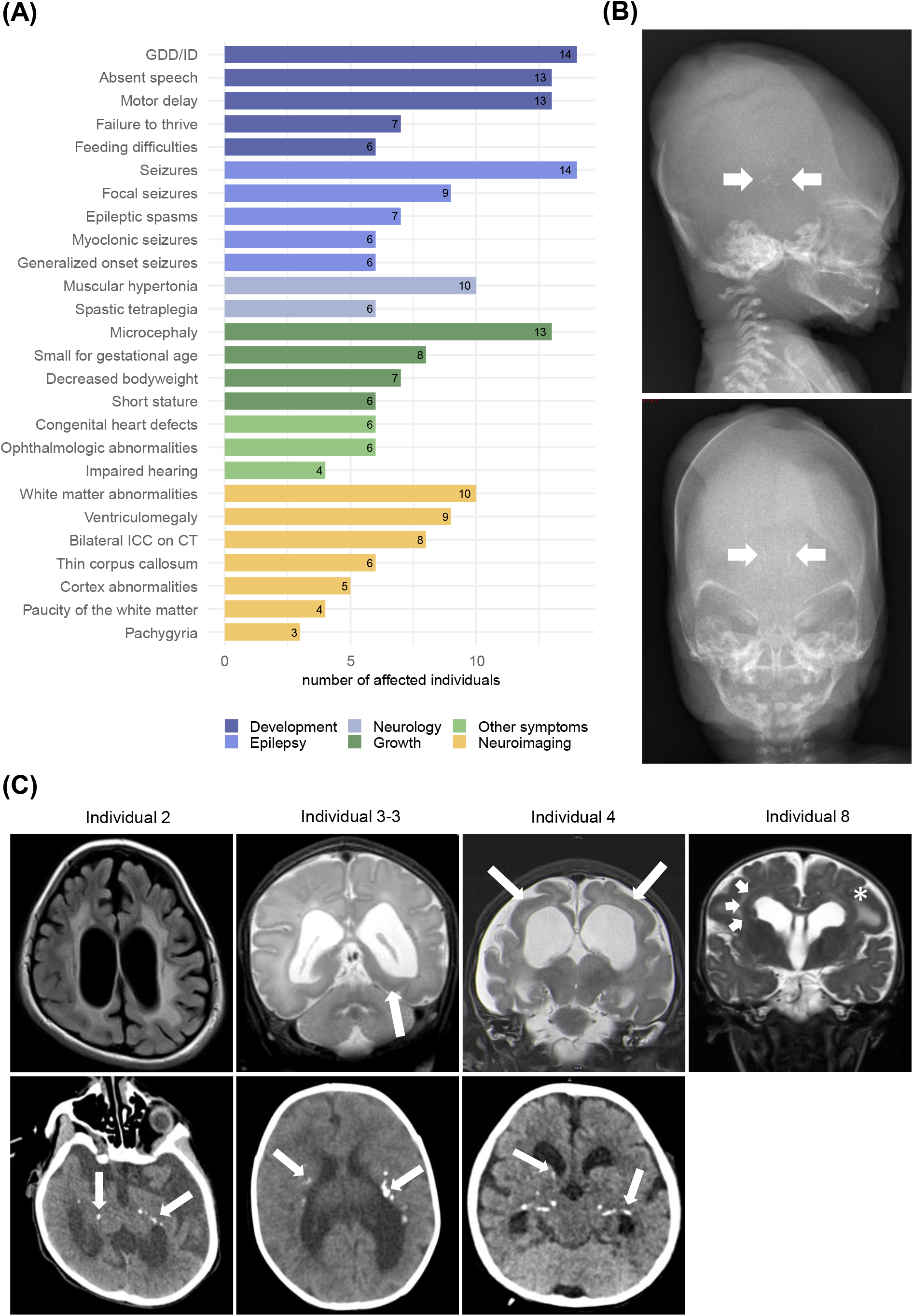
Prevalence of clinical findings, neuroimaging features and X-ray of the fetus. **(A)** Prevalence of phenotypic features in the cohort grouped by clinical categories **(B)** Fetus, (removed due to medRxivpolicy), X-ray babygram postmortem: macroscopic intracranial calcifications (arrows) **(C)** Exemplary MRI and CT images of, each column shows images of one individual as indicated at the top. Ind. 2: a) MRI, age (removed due to medRxivpolicy), T2-FLAIR transverse: pronounced leukoencephalopathy, paucity of the white matter, consecutive ventriculomegaly b) CT, age (removed due to medRxivpolicy): bilateral symmetrical calcifications periventricular and in the basal ganglia (arrows) ; Ind. 3-3: a) MRI, age (removed due to medRxivpolicy), T2-TSE coronal: moderate ventriculomegaly with accentuation of the occipital horn and pachygyria with thickening of the occipitotemporal cortex (arrow) b) CT, (removed due to medRxivpolicy), bilateral calcifications periventricular and in the deep white matter (arrows); Ind. 4: a) MRI, age (removed due to medRxivpolicy), T2-TSE coronal: severe paucity of the white matter and consecutive ventriculomegaly, bilateral pachygyria and thickening of the parietal cortex (arrows) b) CT, age (removed due to medRxivpolicy): bilateral symmetrical calcifications periventricular and in the basal ganglia (arrows); Ind 8: MRI, age (removed due to medRxivpolicy), T2-TSE coronal: ventriculomegaly, pachygyria with thickening of the cortex (asterisk) and periventricular grey matter heterotopia (arrow).

### Neuroimaging

Neuroimaging revealed abnormalities of brain morphology in all twelve individuals that underwent MRI except for individual 1 who had a normal MRI at the age of (removed due to medRxivpolicy). Ten individuals presented signs of leukoencephalopathy (10/12) mainly in a periventricular localisation (8/12) (**Figure 1C**). Four of the individuals showed paucity of the white matter (4/12). For each of the individuals 3-2 and 4, MRI data from two different time points was available, which suggested a progression of the periventricular hyperintensities and loss of white matter, respectively. Five individuals had abnormalities of the cortex morphology (5/12) including pachygyria and thickening of the cortex in three individuals (3/12) (**Figure 1C**) with one of them showing severe periventricular grey matter heterotopia (1/12; **Figure 1C**: Individual 8). Ventriculomegaly of variable degree (8/12) was a common finding. Other notable findings included a thin corpus callosum (6/12), cerebellar vermian hypoplasia (2/12) and a Blake’s pouch cyst in individual 2. Head CT scan, performed in eight individuals, revealed bilateral intracranial calcifications (ICC) in all of them (8/8). Calcifications mostly appeared in a scattered pattern with periventricular localisation (8/8) but also the basal ganglia (4/8), centra semiovale (2/8) and internal capsule (1/8) were affected (**Figure 1C**). Furthermore, CT scan also showed ventriculomegaly in individual 5-2 who did not have MRI.

### Fetal phenotype

The fetus (individual 11) showed severe intrauterine growth retardation and microcephaly during pregnancy and the pregnancy was terminated in the 25^th^ gestational week. Autopsy confirmed length and weight below -2 SD and an occipitofrontal circumference below -4 SD. An x-ray babygram showed ICCs (**Figure 1B**) and in the histopathological examination of the brain, predominant macrocalcification and rare necrotic foci in the process of calcification in the germinative and periventricular areas around the 3^rd^ ventricle and occipital horns were described, as well as cerebral edema with spongiosis and glial response.

### Genetic results

ES revealed homozygous LoF variants in *PPFIBP1* (NM_003622.4) in all affected individuals. In the affected individuals of the families 2, 3, 4, 5, 6, 7, 8, 9 and 10, we detected eight different homozygous protein-truncating variants. These comprise five nonsense variants and three frameshift variants of which one was recurrent in all affected individuals from the two unrelated families 3 and 4 (all variants are displayed in **Table 1**). Since all of these variants lead to premature termination codons >50 nucleotides upstream of the last exon-exon splice junction considering the transcripts with the highest expression overall and in brain tissues in specific (NM_003622 and NM_001198915.2),^14,15^ they are predicted to undergo nonsense mediated mRNA decay (NMD).^29^ Only for the variant c.403C>T, p.(Arg135*), NMD can only be predicted with respect to transcript NM_003622.4, as the variant lies in the 5’ untranslated region (UTR) of the transcript NM_001198915.2. All LoF variants mentioned above can be classified as pathogenic according to the guidelines of the ACMG (Table S1).

In family 1, a homozygous splice-site variant c.1146+1G>A, p.? affecting the consensus 5’-splice site of exon 13 was identified. Multiple *in silico* tools consistently predict a loss of the splice site (Table S2). This could lead to out-of-frame exon skipping or to intron retention.^30,31^ Thus, the mRNA resulting from this allele is likely to encode a truncated protein as well.

Furthermore, in a fetus, a homozygous missense variant c.2177G>T, p.(Gly726Val) was identified. The missense variant lies in the second SAM-domain (**Figure 2A**) and affects a highly conserved amino acid considering nine species up to the Opossum. Multiple *in silico* tools consistently predicted a damaging effect of the variant (Table S3). Structural analysis showed that Gly726 is located in a tight turn of the second SAM-domain of *PPFIPB1* (**Figure 2B**). At this position, a valine can only be accommodated in a strained backbone conformation resulting in domain destabilization. In addition, the longer valine sidechain causes steric problems with Asn803 located in the third SAM domain (**Figure 1B**), which are predicted to disrupt the domain interface.

**Figure 2.**
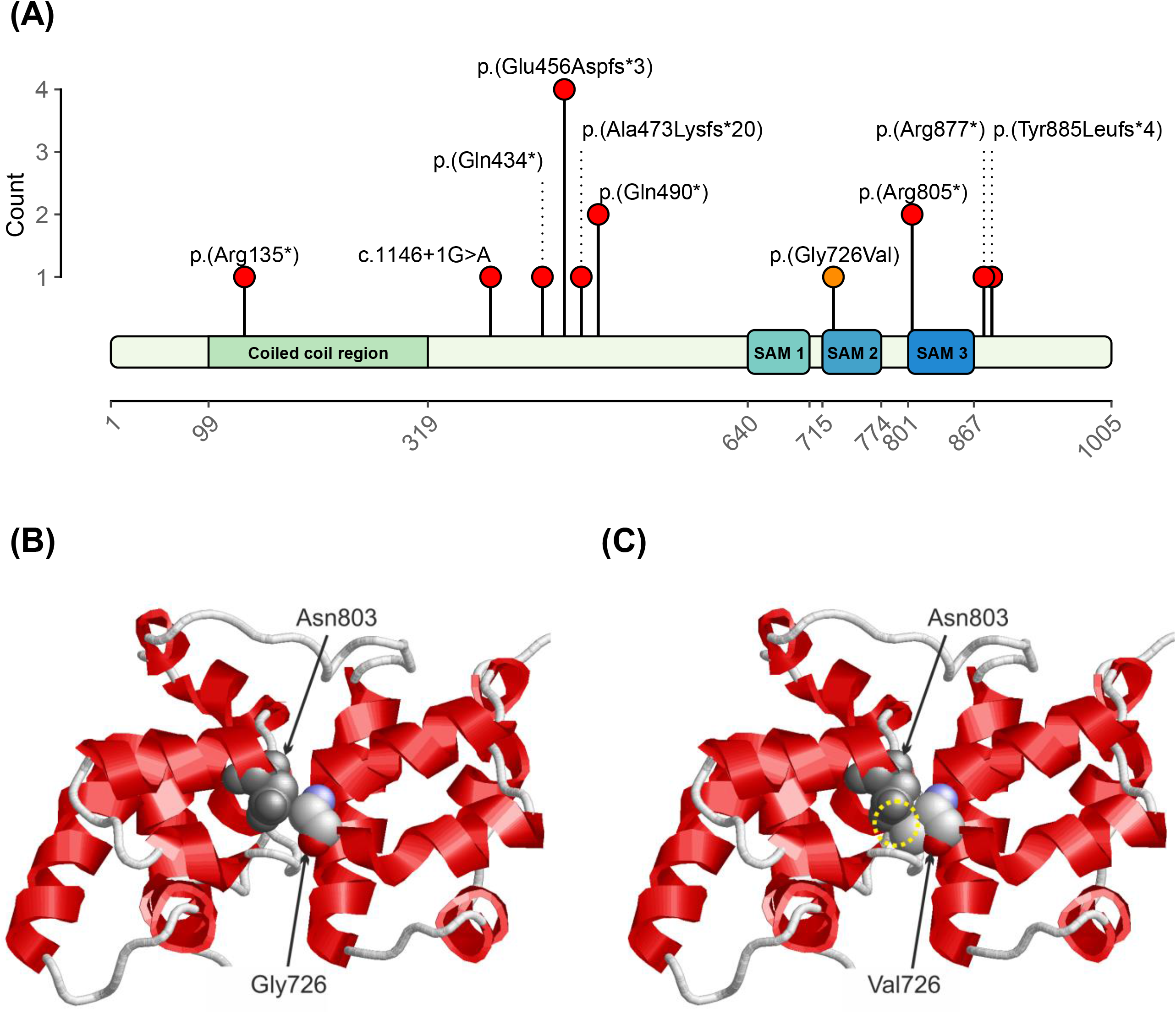
Variant locations on protein level and structure of liprin-β1 illustrating the effect of the p.(Gly726Val) exchange. **(A)** Location of the variants on protein level aligned to the liprin-β1 isoform 1 (GenBank: NP_003613.4 [GenBank: NM_003622.4]). and count of each variant in the cohort. Truncating and splice-site variants are indicated by red dots, the missense variant is indicating by an orange dot. Abbreviation: SAM, sterile alpha motif. **(B)** Structure of the sterile alpha motif (SAM) domains in the wildtype protein. Gly726 is located in a tight turn and makes interactions with Asn803 of the adjacent SAM-domain. Asn803 is shown in grey and Gly726 is colored by atom type; the topology of the protein backbone is schematically depicted with helices in red. **(C)** In the Gly726Val variant, a severe steric overlap (yellow circle) between the sidechains of Val726 and Asn803 is observed, which will disrupt the domain interface thereby altering the topology of the SAM-domain region.

All of the identified variants are very rare in the general population, represented by the gnomAD database.^14^ The variants detected in the families 1, 2, 3, 4, 6, 8, 9, 10 and in the fetus are absent from gnomAD. Five alleles are reported for the variant identified in family 5 (MAF of 0.0000199) and 7 alleles are reported for the variant identified in family 7 (MAF of 0.00002828), all in heterozygous state in each case. The parents were confirmed as heterozygous carriers in the families 1, 2, 4, 5, 6, 7, 8 and 11.

### Modelling loss of *PPFIBP1* in *C. elegans*

Worm models are useful for modelling the underlying mechanistic causes of genetic disorders. CRISPR-Cas9 was used to knock out the *C. elegans PPFIBP1* homolog, *hlb-1*,^10^ and generate a putative LoF mutant, *hlb-1(syb4896)*. Automated quantitative phenotyping of the disease model mutant was then used to identify differences compared to the wild-type strain N2 across a range of behavioural dimensions.^26^

The *hlb-1(syb4896)* mutant showed a significant increase in body curvature (**Figure 3A**). In existing *C. elegans* models of epilepsy “head bobbing” is a phenotype associated with convulsions and the onset of seizures.^32^ We saw no statistically significant difference in the head movement of *hlb-1(syb4896)* compared to N2 during baseline (pre-stimulus) tracking (**Figure 3B**). However, upon stimulation with pulses of blue light, a significant increase in the acceleration of the head tip (indicative of increased head movement) was observed for mutant strains (**Figure 3C**), highlighting some overlap in the behavioural phenotype of *hlb-1(syb4896)* and other pre-existing worm models of epilepsy. Thus, this finding indirectly suggests some elements of a mild epileptic phenotype may be present in *hlb-1(syb4896)*.

**Figure 3.**
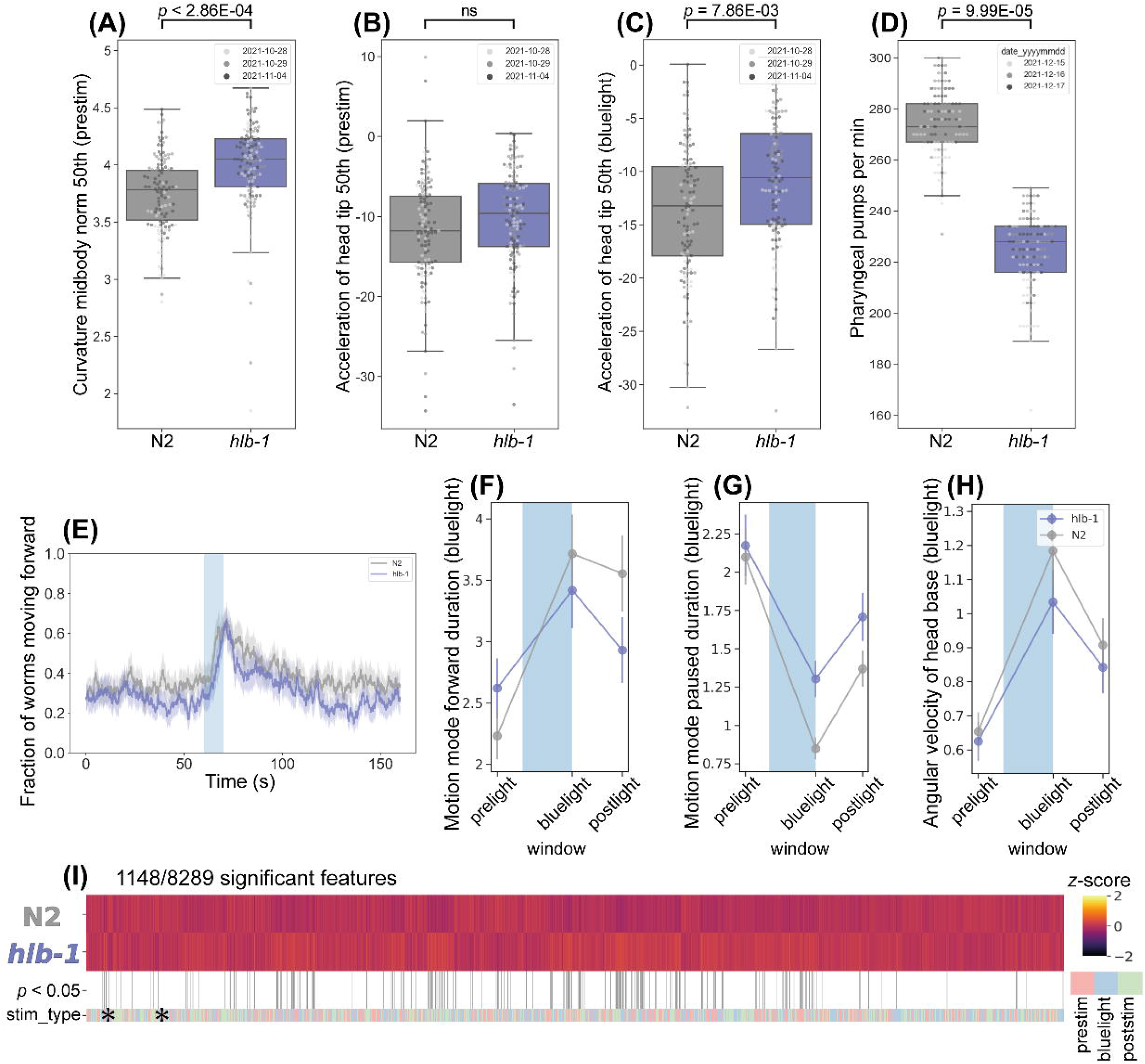
Behavioural phenotype of *Caenorhabditis elegans PPFIBP1* homolog, *hlb-1(syb4896)*. **(A-C)** Example behavioural and postural features altered in the loss-of-function *hlb-1(syb4896)* [*C. elegans* ortholog of *PPFIBP1*] mutant strain under baseline (pre-stimulus) imaging conditions. Individual points marked on the box plots are average values from multiple worms in a single well. The different point colours indicate data from independent experimental days. The selected features were compared to the N2 reference strain using block permutation t-tests and *p*-values are shown above the respective plots. **(D)** Pharyngeal pumps per minute of *hlb-1(syb4896)* and N2 reference strain. **(E)** Overall fraction of worms moving forwards 60 seconds prior and 80 seconds following stimulation with a 10 second blue light pulse (blue shading). Coloured lines represent averages of the detected fraction of paused worms across all biological replicates and shaded areas represent the 95% confidence intervals. **(F-G)** Average changes in the total fraction of worms moving forward or paused prior to, during and following stimulation with blue light, **(H)** average change in an example postural feature in response to blue light. Feature values were calculated as averages of 10 second window summaries centred around 5 seconds before, 10 seconds after and 20 seconds after the beginning of a 10 second blue light pulse (blue shading). **(I)** Heatmap of the entire set of 8289 behavioural features extracted by Tierpsy for *hlb-1(syb4896)* and N2. The stim_type barcode denotes when during image acquisition the feature was extracted: pre-stimulation (pink), blue light stimulation (blue) and post-stimulation (green). Asterisks indicate the selected features present in the box plots above (A-C) and the colour map (right) represents the normalised z-score of the features.

There is little difference in the baseline locomotion of *hlb-1(syb4896)* and N2 (**Figure 3E**). However, *hlb-1(syb4896)* displays a short-lived photophobic escape response when pulsed with blue light, as demonstrated by the LoF mutant returning to a paused state faster upon the cessation of the aversive stimulus (**Figure 3F-G**). We also note that there is an attenuated change in posture of *hlb-1(syb4896)* during blue light tracking (**Figure 3H**).

A previous study into the function of *hlb-1* in *C. elegans* identified a defect in pharyngeal pumping rate,^10^ that we also confirm for *hlb-1(syb4896)* (**Figure 3D**), and enlarged pre- and post-synaptic sites. Given the role of aberrant synaptic transmission events in the onset of epileptic seizures and the hypothesis that liprin-ß1 acts as a core scaffold to mediate protein assembly in the presynaptic zone,^3^ we investigated if our quantitative phenotyping approach could detect a defect in the synaptic transmission apparatus of *hlb-1(syb4896)*.

Aldicarb is an acetylcholinesterase inhibitor that induces paralysis of the body-wall muscles in *C. elegans* due to an accumulation of acetylcholine (ACh) and the subsequent overstimulation of acetylcholine receptors. Increased resistance to aldicarb occurs if mutations give rise to defects in presynaptic function, as ACh accumulates in the neuromuscular junction at a slower rate.^33^ Indeed, *hlb-1(syb4896)* showed a significant dose-dependent decrease in the fraction of paused worms that were exposed to 1 – 10 µM aldicarb for 1h compared to N2 (**Figure 4A**), demonstrating increased aldicarb resistance. Levamisole is a paralysis-inducing ACh receptor agonist. Resistance to levamisole has been shown to persist in worms if mutations affect the postsynaptic site, whereas sensitivity to levamisole persists if mutations only affect the presynaptic site.^34^ In contrast to previous *hlb-1* studies ^10^, we do not observe any resistance to levamisole in *hlb-1(syb4896)* worms. If anything, there is an increased sensitivity observed at 10 µM levamisole for 4h (**Figure 4B**).

**Figure 4.**
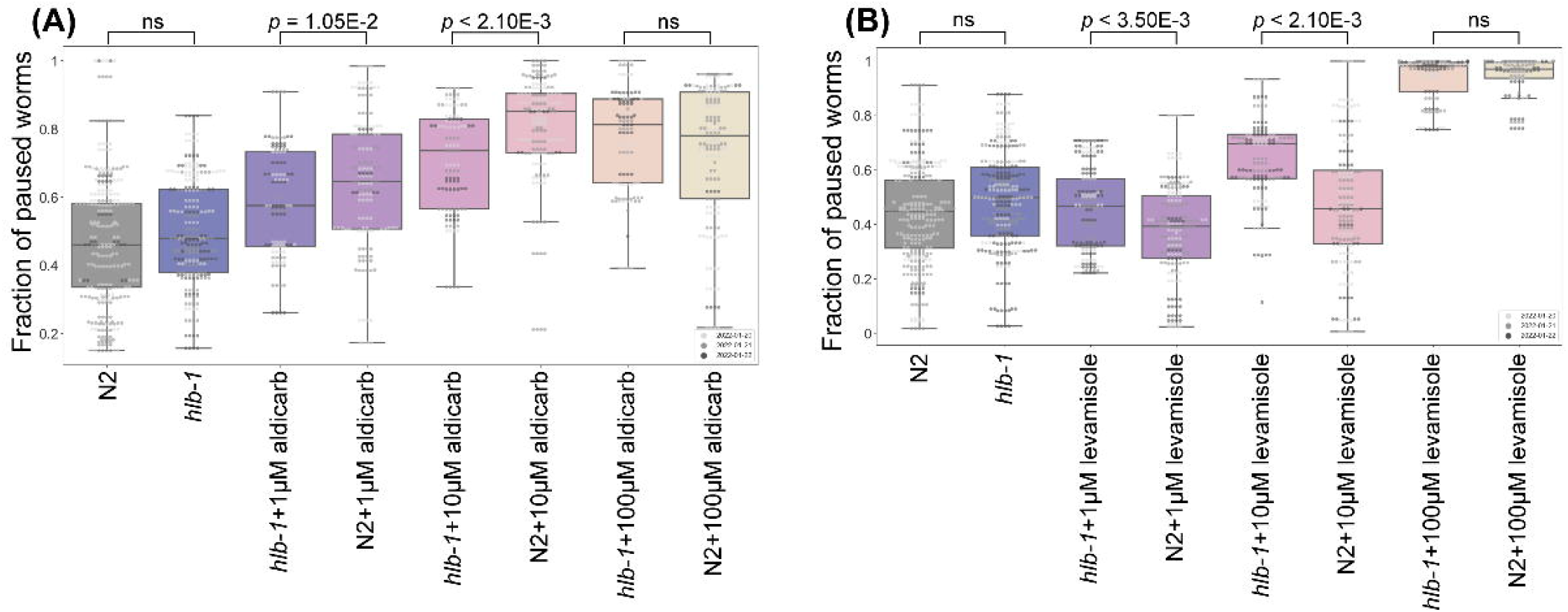
Fraction of paused worms in response to treatment with aldicarb or levamisole. Overall fraction of paused worms after **(A)** 1h exposure to aldicarb and **(B)** 4h exposure to levamisole at the concentrations denoted under the boxplots. N2 (grey) and *hlb-1* (blue) are solvent only controls (DMSO and ddH2O for aldicarb and levamisole, respectively). Individual points marked on the box plots are averaged values from multiple worms in a single well. The different point colours indicate data from independent experimental days. The fraction of paused *hlb-1(syb4896)* worms was compared to the fraction of paused N2 worms at each concentration using block permutation t-tests, with *p* > 0.05 considered not significant (ns), n = 30 wells for each compound and concentration tested.

These findings provide evidence that a defect arises in the presynaptic, but not postsynaptic, apparatus of *C. elegans* due to *hlb-1* LoF. Coupled with existing evidence that liprins are involved in the assembly of presynaptic active zones across species,^3,7^ this points towards a conserved biological role of *hlb-1* and its homologs in regulating the formation of NMJs and supports presynaptic defects as a cause the pathologies arising from mutations in *PPFIBP1*.

## Discussion

Here we describe 14 individuals from ten unrelated families with a core phenotype of moderate to profound developmental delay, progressive microcephaly, epilepsy and periventricular calcifications. In all 14 individuals, ES revealed rare homozygous LoF variants in *PPFIBP1*. In addition, we describe a fetus with severe growth restriction, microcephaly and intracranial calcifications with a homozygous missense variant that is *in silico* and structurally predicted to be disrupting.

Consistent with the proposed autosomal recessive inheritance, LoF variants in *PPFIBP1* in general are not common with an observed/expected ratio (o/e) of 0.57 (90% confidence interval= 0.43-0.75). In addition, there were no homozygous LoF variants observed in gnomAD. Since all described variants are ultra-rare, (MAF < 0.01) it is highly unlikely to assemble a cohort with this level of phenotypical overlap and homozygous LoF variants in *PPFIBP1* by coincidence which further strengthens disease causality.

The 13 individuals harbouring homozygous frameshift or nonsense variants exhibit a consistent phenotype in terms of the severity of the developmental delay, epilepsy and frequently found neuroimaging features. Only individual 7 presented a milder disease course compared to the other individuals with truncating variants as she had secondary microcephaly, was able to stand and walk, albeit showing impaired motor development, and showed less prominent neuroimaging features. The nonsense variant c.403C>T, p.(Arg135*) found in individual 7 lies in the 6^th^ exon thus being the most upstream variant in this cohort. It is predicted to cause NMD considering the transcript NM_003622.4 which shows the highest overall expression and thereby likely has the highest biological relevance. Nevertheless, it cannot be ruled out that a shorter transcript like NM_001198915.2 with its start codon lying 57 base pairs downstream of this variant could compensate for the loss of the main transcript to some extent. NM_001198915.2 has the second highest mean expression across all tissues and particularly shows expression levels that are comparable to those of NM_003622.4 in some areas of the brain.^15^

Individual 1 with the homozygous splice-site variant has a milder phenotype compared to the other individuals with nonsense or frameshift variants, although he shares the core clinical signs. This could be due to an incomplete splice defect, either leading to the expression of a fraction of normal protein or to an altered protein not completely impaired in function or stability. Canonical splice site variants as observed in individual 1 can have a variety of effects on pre-mRNA splicing such as exon skipping which is the most common mechanism in variants disrupting consensus 5’-donor splice sites^30^ or intron retention, both of which would result in a frameshift in this case. However, a loss of the splice site could also enable the activation of a cryptic splice site with consequent inclusion of an intron fragment or the removal of an exon fragment, both of which can lead to a variety of aberrant transcripts.

The pathogenicity of the missense variant identified in the fetus is not as clear as that of the LoF variants. However, the striking similarity of the intracranial calcifications, the growth restriction and the severe microcephaly represent a significant phenotypic overlap with the rest of the cohort, suggesting this variant to be causative. Potential pathogenicity of the variant is further supported by its absence from the general population, multiple supported by *in silico* predictions and its expected effects on the SAM-domains from structural analysis. SAM-domains are a family of protein interaction modules present in a wide variety of proteins.^35^ The Gly726Val exchange is located in the second SAM-domain of PPFIBP1 destabilizing both the second SAM-domain and the interaction between the second and third SAM domain. Therefore, this variant is expected to severely disturb the topology of the SAM-domain region and its function in protein-protein interactions. Given the hypothesis that liprin-β1 acts as a core scaffold to mediate protein assembly in the presynaptic zone,^3^ the ability to precisely interact with other proteins would appear to be critical for protein function.

ICCs located in the periventricular area but also affecting the basal ganglia and the internal capsule appear to be a highly characteristic sign in this cohort. Pathologic ICCs have heterogeneous aetiologies such as neoplastic, infectious, vascular, metabolic and genetic conditions.^36^ Congenital infections with pathogens of the TORCH-spectrum, and congenital cytomegalovirus (CMV) infections in particular, account for a significant amount of congenital and paediatric ICCs that are associated with brain malformations and impaired neurodevelopment.^37^ However, genetic disorders such as interferonopathies represent important differential diagnoses for congenital ICC and some conditions significantly overlap with the symptomatic spectrum of congenital TORCH-infections.^38–40^ It is assumed that the genetic aetiologies of unsolved ICCs have not been fully discovered yet.^37,41^ Individual 4 was admitted to the neonatal intensive care unit for one month after birth, as his clinical presentation was indicative of a congenital CMV infection (see case reports in Supplemental data for further details). However, an active CMV infection could not be confirmed in standard laboratory diagnostics. In both affected siblings of family 5 and in individual 6-2, a screening for infections of the TORCH spectrum was performed with negative results and also the fetus was tested negative for CMV.

The biological function of liprin-β1 and its molecular mechanisms are still largely unstudied. However, recent studies point towards a role in neurodevelopment that echo the findings of a neurodevelopmental disorder in the cohort described here. Liprin-β1 has been identified as a binding partner of liprin-α proteins. The role of liprin-α proteins or their orthologues, in synapse formation and synaptic transmission has been demonstrated in previous animal model studies.^7,42–44^ Liprin-α proteins function as major scaffold proteins at the presynaptic active zone and at the postsynaptic density and also play a role in intracellular transport, cell motility and protein assembly.^3,5,6,45–47^ Wei *et al*. found that liprin-α2 forms a ternary complex simultaneously binding liprin-β1 and CASK, another presynaptic scaffold protein, supporting the hypothesis that liprin-β1 could act as a core scaffold and mediate large protein assemblies in the presynaptic active zone.^3^ Interestingly, pathogenic variants in *CASK* (MIM: 300172) are associated with X-linked neurodevelopmental disorders.^48^ Pathogenic variants in *CASK* cause X-linked neurodevelopmental disorders with varying phenotypes depending on variant type and inheritance. In particular, heterozygous and hemizygous LoF variants in *CASK* lead to Microcephaly with pontine and cerebellar hypoplasia (MICPCH [MIM: 300749]). The phenotypic spectrum comprises moderate to profound ID, progressive microcephaly, impaired hearing, ophthalmologic anomalies, muscular hypo- or hypertonia and spasticity, as well as seizures and partly epileptic encephalopathy in males.^48^ Since the phenotype is overlapping with the clinical signs found in this cohort, it seems possible that *PPFIBP1* and *CASK* are involved in similar biological functions such as protein assembly in the presynaptic active zone. Supporting the potential role of liprin-β1 in synapse formation and neurodevelopment, we have shown here that a *C. elegans PPFIBP1/hlb-1* knockout model shows defects in spontaneous and light-induced behaviour. The observed sensitivity of the worm model to the acetylcholinesterase inhibitor aldicarb supports a presynaptic defect as at least a partial cause of the observed behavioural phenotypes. This is broadly consistent with previous work showing that null-allele mutants of the drosophila orthologues liprin-β and liprin-α independently cause abnormal axon outgrowth, target layer recognition and synapse formation of R7 photoreceptors as well as reduced larval NMJ size in *Drosophila melanogaster*. Interestingly, distinct effects on axon outgrowth between single liprin-β and liprin-α mutants and an additive effect in double mutants were observed, indicating independent functions of both proteins.^7^

In summary, we establish bi-allelic loss-of-function variants in *PPFIBP1* as a novel cause for an autosomal recessive neurodevelopmental disorder with early-onset epilepsy, microcephaly and periventricular calcifications.

## Data Availability

The code used for tracking and extracting C. elegans behavioural features is available at https://github.com/Tierpsy and code for performing statistical analysis and generating figures is available at https://github.com/Tom-OBrien/Phenotyping-hlb1-disease-model-mutant. The associated C. elegans datasets are available at https://doi.org/10.5281/zenodo.6338403. The Morbid Genes Panel is available here https://morbidgenes.org/ and here https://zenodo.org/record/6136995#.YiYvI-jMKUk. Protocols used in this study are available here: Barlow, Ida. Bleach Synchronisation of C. Elegans V1. https://doi.org/10.17504/protocols.io.2bzgap6. Barlow, Ida. Disease Model Screen Protocol V1. https://doi.org/10.17504/protocols.io.bsicncaw. Islam, Priota. Preparing Worms for the COPAS (Wormsorter) V1. https://doi.org/10.17504/protocols.io.bfqbjmsn. J OBrien, Thomas. Pharyngeal Pumping Assay V1.
https://doi.org/10.17504/protocols.io.b3hiqj4e. J. OBrien, Thomas. Response of Disease Model Mutants to Cholinergic Drugs V1. https://doi.org/10.17504/protocols.io.b5p7q5rn. Moore, Saul, and Ida Barlow. COPAS Wormsorter V1.
https://doi.org/10.17504/protocols.io.bfc9jiz6. Barlow, Ida. Disease Model Screen Protocol V1. https://doi.org/10.17504/protocols.io.bsicncaw.

https://morbidgenes.org/

https://zenodo.org/record/6136995#.YiYvI-jMKUk

https://doi.org/10.17504/protocols.io.2bzgap6.

https://doi.org/10.17504/protocols.io.bsicncaw.

https://doi.org/10.17504/protocols.io.bfqbjmsn.

https://doi.org/10.17504/protocols.io.b3hiqj4e.

https://doi.org/10.17504/protocols.io.b5p7q5rn.

https://doi.org/10.17504/protocols.io.bfc9jiz6.

https://doi.org/10.17504/protocols.io.bsicncaw.

## Data and code availability

The code used for tracking and extracting *C. elegans* behavioural features is available at https://github.com/Tierpsy and code for performing statistical analysis and generating figures is available at https://github.com/Tom-OBrien/Phenotyping-hlb1-disease-model-mutant. The associated *C. elegans* datasets are available at https://doi.org/10.5281/zenodo.6338403.

The Morbid Genes Panel is available here https://morbidgenes.org/ and here https://zenodo.org/record/6136995#.YiYvI-jMKUk

Protocols used in this study are available here:

Barlow, Ida. Bleach Synchronisation of C. Elegans V1.

https://doi.org/10.17504/protocols.io.2bzgap6.

Barlow, Ida. Disease Model Screen Protocol V1.

https://doi.org/10.17504/protocols.io.bsicncaw.

Islam, Priota. Preparing Worms for the COPAS (Wormsorter) V1.

https://doi.org/10.17504/protocols.io.bfqbjmsn.

J O’Brien, Thomas. Pharyngeal Pumping Assay V1.

https://doi.org/10.17504/protocols.io.b3hiqj4e.

J. O’Brien, Thomas. Response of Disease Model Mutants to Cholinergic Drugs V1.

https://doi.org/10.17504/protocols.io.b5p7q5rn.

Moore, Saul, and Ida Barlow. COPAS Wormsorter V1.

https://doi.org/10.17504/protocols.io.bfc9jiz6.

Barlow, Ida. Disease Model Screen Protocol V1.

https://doi.org/10.17504/protocols.io.bsicncaw.

## Supplemental information

Supplemental data includes detailed case reports of the described individuals, three tables (Table S1, Table S2, Table S3) and supplemental methods. Table S4 is provided separately as an excel file.

## Acknowledgements

We thank all families that participated in this study. This project has received funding from the European Research Council (ERC) under the European Union’s Horizon 2020 Research and Innovation Program (Grant Agreement No. 714853) and was supported by the Medical Research Council through Grant MC-A658-5TY30. HT was supported by the European Union’s Seventh Framework Programme for research, technological development and demonstration under grant agreement no. 608473.

## Web resources

GenBank, https://www.ncbi.nlm.nih.gov/genbank/

OMIM, https://www.omim.org/

GeneMatcher, https://genematcher.org/

MorbidGenes, https://morbidgenes.org/

gnomAD, https://gnomad.broadinstitute.org/

GTEx, https://gtexportal.org/home/

Ensembl, https://www.ensembl.org/index.html

PDB, https://www.rcsb.org/

SwissModel, https://swissmodel.expasy.org/

