## Supplemental Data for "Bi-allelic loss-of-function variants in *PPFIBP1* cause a neurodevelopmental disorder with microcephaly, epilepsy and periventricular calcifications"

### Inhalt

### Supplemental case reports

#### *Individual 1:*

This case report has been removed from the preprint version to comply with medrxiv policy. Please see the published version or contact the authors if you are interested in this information.

#### *Individual 2:*

This case report has been removed from the preprint version to comply with medrxiv policy. Please see the published version or contact the authors if you are interested in this information.

#### *Individual 3-1, 3-2 and 3-3:*

This case report has been removed from the preprint version to comply with medrxiv policy. Please see the published version or contact the authors if you are interested in this information.

#### *Individual 4:*

This case report has been removed from the preprint version to comply with medrxiv policy. Please see the published version or contact the authors if you are interested in this information.

#### *Individual 5-1 and 5-2:*

This case report has been removed from the preprint version to comply with medrxiv policy. Please see the published version or contact the authors if you are interested in this information.

#### *Individual 6-1 and 6-2*

Individual 6-1:

This case report has been removed from the preprint version to comply with medrxiv policy. Please see the published version or contact the authors if you are interested in this information.

*Individual 7*

This case report has been removed from the preprint version to comply with medrxiv policy. Please see the published version or contact the authors if you are interested in this information.

*Individual 8*

This case report has been removed from the preprint version to comply with medrxiv policy. Please see the published version or contact the authors if you are interested in this information.

*Individual 9*

This case report has been removed from the preprint version to comply with medrxiv policy. Please see the published version or contact the authors if you are interested in this information.

*Individual 10*

This case report has been removed from the preprint version to comply with medrxiv policy. Please see the published version or contact the authors if you are interested in this information.

*Fetus (family 11)*

The clinical and molecular findings in the fetus are fully described in the main article.

**Table S1.** Variant information and classification according to the ACMG criteria[1,*]. Variant descriptions were validated according to the HGVS nomenclature.

| Individual | Chr | Genomic position (GRCh38) | cDNA (NM_003622.4) | Protein (NP_003613.4) | Allelic state | Predicted effect | ACMG criteria | Classification |
|---|---|---|---|---|---|---|---|---|
| 1 | 12 | g.27667321G>A | c.1146+1G>A | p.? | homozygous | Loss of 5'-donor splice site of exon 13 | PVS1, PM2_Supporting, PM3_Supporting | Pathogenic |
| 2 | 12 | g.27689172del | c.2654del | p.(Tyr885Leufs*4) | homozygous | Nonsense mediated mRNA decay | PVS1, PM2_Supporting, PM3_Supporting | Pathogenic |
| 3-1, 3-2, 3-3, 4 | 12 | g.27673815_27673816del | c.1368_1369del | p.(Glu456Aspfs*3) | homozygous | Nonsense mediated mRNA decay | PVS1, PM2_Supporting, PM3_Supporting, PP1_Moderate | Pathogenic |
| 5-1, 5-2 | 12 | g.27688340C>T | c.2413C>T | p.(Arg805*) | homozygous | Nonsense mediated mRNA decay | PVS1, PM2_Supporting, PM3_Supporting | Pathogenic |
| 6-1, 6-2 | 12 | g.27676485C>T | c.1468C>T | p.(Gln490*) | homozygous | Nonsense mediated mRNA decay | PVS1, PM2_Supporting, PM3_Supporting | Pathogenic |
| 7 | 12 | g.27647774C>T | c.403C>T | p.(Arg135*) | homozygous | Nonsense mediated mRNA decay | PVS1, PM2_Supporting, PM3_Supporting | Pathogenic |
| 8 | 12 | g.27676434_27676444del | c.1417_1427del | p.(Ala473Lysfs*20) | homozygous | Nonsense mediated mRNA decay | PVS1, PM2_Supporting, PM3_Supporting | Pathogenic |
| 9 | 12 | g.27672464C>T | c.1300C>T | p.(Gln434*) | homozygous | Nonsense mediated mRNA decay | PVS1, PM2_Supporting, PM3_Supporting | Pathogenic |

| | | | | | | | | |
|---|---|---|---|---|---|---|---|---|
| 10 | 12 | g.27689147C>T | c.2629C>T | p.(Arg877*) | homozygous | Nonsense mediated mRNA decay | PVS1, PM2_Supporting, PM3_Supporting | Pathogenic |
| Fetus | 12 | g.27682633G>T | c.2177G>T | p.(Gly726Val) | homozygous | Missense change predicted to severely disturb topology of the SAM-domain region | PM2_Supporting PM3_Supporting PP3 | Uncertain significance ** |

* Although ACMG criteria are formally not perfectly suited for the description of novel disease-associated genes, we classified all variants from this study to simplify further use of the data.

** Due to the high clinical overlap to the rest of the cohort, this variant is deemed causative despite being classified as uncertain.

**Table S2.** *In silico* prediction of the splice variant *PPFIBP1* (NM_003622.4).

| **Ind.** | **Genomic position** (GRCh38) | **cDNA** | **CADD-v6**[2] | **SpliceAI**[3] | **MaxEntScan**[4] | **NNSPLICE**[5] | **Nucleotide conservation** | **gnomAD**[6] |
|---|---|---|---|---|---|---|---|---|
| 1 | chr12:g.27667321 | c.1146+1G>A, p.? | 35 | 0.95 | 1 | 1 | High | 0 |

Red color represents a very high probability of the variant to be damaging for each aspect in the table.

**Table S3.** *In silico* prediction of the missense variant and conservation of the affected amino acid in *PPFIBP1*(NM_003622.4).

| **Ind.** | **Genomic position** (GRCh38) | **cDNA** | **CADD-v6**[2] | **REVEL**[7] | **Mutation Taster**[8] | **M-CAP 1.3**[9] | **Polyphen 2 v2.2.2**[10] | **GERP++**[11] | **AA conservation** | **gnomAD**[6] |
|---|---|---|---|---|---|---|---|---|---|---|
| Fetus (11) | chr12:27682633 | c.2177G>T, p.(Gly726Val) | 24.6 | 0,609 (LDC) | D | 0.063 (PoP) | PrD | 4,46 | High | 0 |

Red color represents a very high probability of the variant to be damaging for each aspect in the table; LDC = likely disease causing; D = Deleterious; PoP = Possibly Pathogenic; PrD = Probably Damaging

### Supplemental Methods

#### *Individual 1*

Trio exome sequencing of the index and the parents was performed at the Institute of Human Genetics at the University of Leipzig Medical Center. Library preparation was done using the Nextera DNA Flex Pre-Enrichment LibraryPrep with Illumina Nextera DNA UD Indexes by Illumina (San Diego, CA, USA). Target enrichment was achieved by using the Human Core Exome hybridization probes from Twist Bioscience (San Francisco, CA, USA). Paired-end Next-Generation-Sequencing was then performed on a NovaSeq 6000 Instrument (Illumina), located at the facilities of GeneWIZ (Leipzig, Germany), using an S1 Reagent Kit (300 cycles) by Illumina. Coverage of more than 20x has been achieved in more than 95 % of target sequences in all family members.

Analysis of the raw data, variant annotation and prioritization were performed using the software Varfeed (Limbus, Rostock, Germany) and Varvis (Limbus, Rostock, Germany). Variants were prioritized based on the mode of inheritance, impact on the gene product, minor allele frequency and *in silico* predicted pathogenicity. For research evaluation of variants in potential candidate genes, we also considered the biological function of the gene product with regards to neurodevelopment, mutational constraint parameters[6] (i.e. observed/expected - ratio, pLI-Score, Z-Score), expression patterns of the gene[12], insights on animal models and further aspects such as protein interaction networks and the function of similar or related proteins.

#### *Individual 2*

Trio-like exome sequencing of the index case and the parents was performed as previously described[13]. Library preparation was done using the Nextera DNA Flex Pre-Enrichment LibraryPrep with Illumina Nextera DNA UD Indexes by Illumina (San Diego, CA, USA). Target enrichment was achieved by using a modified version of the Human Core Exome hybridization probes from Twist Bioscience (San Francisco, CA, USA) complemented with additional custom probes. Paired-end Next-Generation-

Sequencing (2x100 bp) was then performed on a NovaSeq 6000 Instrument (Illumina), at IntegraGen SA (Evry, France). Coverage of more than 20X has been achieved in more than 97 % of target sequences in the index case.

Analysis of the raw data, variant annotation and prioritization were performed as previously described [13].

#### *Individuals 3-1, 3-2 and 3-3*

Exome sequencing and variant prioritization supported by autozygome analysis were performed as previously described[14,15].

#### *Individual 4*

Whole exome sequencing of the index and targeted sequencing on both DNA strands of the relevant *PPFIBP1* region for the parents was performed at CENTOGENE (the rare disease company) (Rostock, Germany). Double stranded DNA capture baits against approximately 36.5 Mb of the human coding exome (targeting >98% of the coding RefSeq from the human genome build GRCh37/hg19) are used to enrich target regions from fragmented genomic DNA with the Twist Human Core Exome Plus kit by Twist Bioscience (San Francisco, CA, USA). The generated library is sequenced on an Illumina platform (San Diego, CA, USA) to obtain at least 20x coverage depth for >98% of the targeted bases. An in-house bioinformatics pipeline, including read alignment to GRCh37/hg19 genome assembly, variant calling, annotation and comprehensive variant filtering is applied. All variants with minor allele frequency (MAF) of less than 1% in gnomAD[6] database, and disease-causing variants reported in HGMD[16], in ClinVar[17] or in CentoMD® by CENTOGENE are considered. The investigation for relevant variants is focused on coding exons and flanking +/-20 intronic nucleotides of genes with clear gene-phenotype evidence (based on OMIM® information). All potential modes of inheritance patterns are considered. In addition, provided family history and clinical information are used to evaluate identified variants with respect to their pathogenicity and causality. Variants are categorized into five classes (pathogenic; likely pathogenic; VUS; likely benign; benign). All variants related to the phenotype of the patient are reported. CENTOGENE has established stringent quality criteria and validation processes for variants detected by next-generation sequencing. Variants with low quality and/or unclear zygosity are confirmed by orthogonal methods. Consequently, a specificity of >99.9% for all reported variants is warranted.

##### *Individuals 5-1 and 5-2*

Genomic DNA was extracted from peripheral blood samples according to standard procedures of phenol chloroform extraction. WES on each proband was performed as described elsewhere[18,19] in Macrogen, Korea. Briefly, target enrichment was performed with 2 μg genomic DNA using the SureSelectXT Human All Exon Kit version 6 (Agilent Technologies, SantaClara, CA, USA) to generate barcoded whole-exome sequencing libraries. Libraries were sequenced on the HiSeqX platform (Illumina, San Diego, CA, USA) with 50x coverage. Quality assessment of the sequence reads was performed by generating QC statistics with FastQC (http://www.bioinformatics.bbsrc.ac.uk/projects/fastqc). Our bioinformatics filtering strategy included screening for only exonic and donor/acceptor splicing variants. In accordance with the pedigree and phenotype, priority was given to rare variants (<0.01% in public databases, including 1,000 Genomes project, NHLBI Exome Variant Server, Complete Genomics 69, and Exome Aggregation Consortium [ExAC v0.2]) that were fitting a recessive (homozygous or compound heterozygous) or a de novo model and/or variants in genes previously linked to developmental delay, intellectual disability, and other neurological disorders.

##### *Individuals 6-1 and 6-2*

Whole-exome sequencing (WES) of the proband was performed using a TruSeq DNA PCR-free library preparation method by Illumina (San Diego, CA, USA) and subsequent sequencing on a NovaSeq 6000 sequencing instrument (Illumina) to a sequencing depth of 30x median coverage. The resulting WES data was analysed using the Mutation Identification Pipeline (MIP) as previously described[20].

Segregation of the variant identified in *PPFIBP1* was carried out by PCR amplification of genomic DNA and subsequent Sanger sequencing using the BigDye version 3.1 sequencing kit (Applied Biosystems, Foster City, CA, USA) on a 3500 Genetic Analyzer (Applied Biosystems).

##### *Individual 7*

Whole-genome sequencing (WGS) of the proband and her parents was performed at the Science for Life Laboratory, Clinical Genomics facility, Stockholm, using a TruSeq DNA PCR-free library preparation method by Illumina (San Diego, CA, USA) and subsequent sequencing on a NovaSeq 6000 sequencing instrument (Illumina) to a sequencing depth of 30x median coverage. The resulting WGS data was analyzed using the Mutation Identification Pipeline (MIP) as previously described[20]. Confirmation of the variant observed in *PPFIBP1* was carried out by PCR amplification of genomic DNA and subsequent Sanger sequencing using the BigDye version 3.1 sequencing kit (Applied Biosystems, Waltham, Massachusetts, USA) on a 3500xl Genetic Analyzer (Applied Biosystems).

##### *Individual 8*

Genomic DNA was extracted from peripheral blood samples according to standard procedures of phenol chloroform extraction. WES on each proband was performed as described elsewhere[18,19] in Macrogen, Korea. Briefly, target enrichment was performed with 2 µg genomic DNA using the SureSelectXT Human All Exon Kit version 6 (Agilent Technologies, SantaClara, CA, USA) to generate barcoded whole-exome sequencing libraries. Libraries were sequenced on the HiSeqX platform (Illumina, San Diego, CA, USA) with 50x coverage. Quality assessment of the sequence reads was performed by generating QC statistics with FastQC (http://www.bioinformatics.bbsrc.ac.uk/projects/fastqc). Our bioinformatics filtering strategy included screening for only exonic and donor/acceptor splicing variants. In accordance with the pedigree and phenotype, priority was given to rare variants (<0.01% in public databases, including 1,000 Genomes project, NHLBI Exome Variant Server, Complete Genomics 69, and Exome Aggregation Consortium [ExAC v0.2]) that were fitting a recessive (homozygous or compound heterozygous) or a de novo model and/or variants in genes previously linked to developmental delay, intellectual disability, and other neurological disorders.

##### *Individual 9*

Exome sequencing of the patient was performed at the Macrogen company (Seoul, South Korea) using Agilent SureSelect V6 post capture kit with a NovaSeq 6000 instrument by Illumina (San Diego, CA, USA). Target regions have a mean read depth of 100X.

Analysis of the raw data was performed using BWA and GATK packages. Variant annotation and prioritization were done using an in-house pipeline of the Palindrome lab; briefly this pipeline uses an ACMG based algorithm and an in-house database for variant prioritization. Clinically significant variants are curated manually.

##### *Individual 10*

Genomic DNA was extracted from peripheral blood samples according to standard procedures of phenol chloroform extraction. WES on each proband was performed as described elsewhere[18,19] in Macrogen, Korea. Briefly, target enrichment was performed with 2 μg genomic DNA using the SureSelectXT Human All Exon Kit version 6 (Agilent Technologies, SantaClara, CA, USA) to generate barcoded whole-exome sequencing libraries. Libraries were sequenced on the HiSeqX platform (Illumina, San Diego, CA, USA) with 50x coverage. Quality assessment of the sequence reads was performed by generating QC statistics with FastQC (http://www.bioinformatics.bbsrc.ac.uk/projects/fastqc). Our bioinformatics filtering strategy included screening for only exonic and donor/acceptor splicing variants. In accordance with the pedigree and phenotype, priority was given to rare variants (<0.01% in public databases, including 1,000 Genomes project, NHLBI Exome Variant Server, Complete Genomics 69, and Exome Aggregation Consortium [ExAC v0.2]) that were fitting a recessive (homozygous or compound heterozygous) or a de novo model and/or variants in genes previously linked to developmental delay, intellectual disability, and other neurological disorders.

#### *Fetus (Family 11)*

Trio exome sequencing of the index and the parents was performed at Laboratoire Biomnis-Eurofins (Lyon, France). Library preparation was done using the Twist Library Preparation from Twist Bioscience (San Francisco, CA, USA). Target enrichment was achieved by using the Twist Human Core Exome hybridization probes from Twist Bioscience. Paired-end Next-Generation-Sequencing was then performed on a NextSeq500 Instrument, using an a NextSeq 500/550 High Output Kit v2 (2x75pb, paired-end) by Illumina (San Diego, CA, USA). Coverage of more than 30x has been achieved in more than 97 % of target sequences in all family members.

Analysis of the raw data, variant annotation and prioritization were performed using the software SeqOne (Montpellier, France).

Sanger verification has been performed with the BigDye v3.1 kit followed by a purification with the BigDye X-terminator from Life Technologies. The electrophoresis has been done on the SeqStudio instrument from Applied Biosystems (Waltham, Massachusetts, USA).
